# Online health search via multi-dimensional information quality assessment based on deep language models

**DOI:** 10.1101/2023.04.11.22281038

**Authors:** Boya Zhang, Nona Naderi, Rahul Mishra, Douglas Teodoro

**Author notes:** **Email(s):** (B. Zhang); (N. Naderi); (R. Mishra); (D. Teodoro).

## Abstract

**Background:** Widespread misinformation in Web resources can lead to serious implications for individuals seeking health advice. Despite that, information retrieval models are often focused only on the query-document relevance dimension to rank results.

**Objective:** We investigate a multi-dimensional information quality retrieval model based on deep learning to enhance the effectiveness of online healthcare information search results.

**Methods:** In this study, we simulated online health information search scenarios with a topic set of 32 different health-related inquiries and a corpus containing one billion Web documents from the April 2019 snapshot of Common Crawl. Using state-of-the-art pre-trained language models, we assessed the quality of the retrieved documents according to their usefulness, supportiveness, and credibility dimensions for a given search query on 6,030 human-annotated query-document pairs. We evaluated this approach using transfer learning and more specific domain adaptation techniques.

**Results:** In the transfer learning setting, the usefulness model provided the largest distinction between help- and harm-compatible documents with a difference of +5.6%, leading to a majority of helpful documents in the top-10 retrieved. The supportiveness model achieved the best harm compatibility (+2.4%), while the combination of usefulness, supportiveness, and credibility models achieved the largest distinction between help- and harm-compatibility on helpful topics (+16.9%). In the domain adaptation setting, the linear combination of different models showed robust performance with help-harm compatibility above +4.4% for all dimensions and going as high as +6.8%.

**Conclusions:** These results suggest that integrating automatic ranking models created for specific information quality dimensions can increase the effectiveness of health-related information retrieval. Thus, our approach could be used to enhance searches made by individuals seeking online health information.

## Introduction

In today’s digital age, individuals with diverse information needs, medical knowledge, and linguistic skills [1] turn to the Web for health advice and make treatment decisions [2]. The mixture of facts and rumors in online resources [3] makes it challenging for users to discern accurate content [4]. To provide high-quality resources and enable properly informed decision-making [5], information retrieval systems should differentiate between accurate and misinforming content [6]. Nevertheless, search engines rank documents mainly by their relevance to the search query [7], neglecting several health information quality concerns. Moreover, despite attempts by some search engines to combat misinformation [8], they lack transparency in terms of the methodology used and performance evaluation.

*Health misinformation* is defined as health-related information that is inaccurate or misleading based on current scientific evidence [9,10]. Due to the lack of health literacy of non-professionals [11] and the rise of infodemic phenomenon [12] — the rapid spread of both accurate and inaccurate information about a medical topic on the internet [13] — health misinformation has become increasingly prevalent online. Topics related to misinformation, such as ‘vaccine’ or ‘the relationship between coronavirus and 5G’ have gained scientific interest across social media platforms like Twitter and Instagram [14–16] and among various countries [17]. Thus, the development of new credibility-centered search methods and assessment measures is crucial to address the pressing challenges in health-related information retrieval [18].

In recent years, numerous approaches have been introduced in the literature to categorize and assess misinformation according to multiple dimensions. Soprano *et al.* [19] proposed seven dimensions of *truthfulness*, which include *correctness, neutrality, comprehensibility, precision, completeness, speaker trustworthiness*, and *informativeness*. On the other hand, Linden *et al.* [20] categorized infodemic into three key dimensions: *susceptibility*, *spread*, and *immunization*. Information retrieval shared tasks, such as the Text REtrieval Conference (TREC) and the Conference and Labs of the Evaluation Forum (CLEF), have also started evaluating quality-based systems for health corpora using multiple dimensions [21,22]. The CLEF eHealth Lab Series proposed a benchmark to evaluate models according to the *relevance*, *readability*, and *credibility* of the retrieved information [23]. The TREC Health Misinformation Track 2021 proposed further metrics *usefulness*, *supportiveness* and *credibility* [24]. These dimensions also appear in the TREC Health Misinformation Track 2019 as *relevancy*, *efficacy*, and *credibility*, respectively. Additionally, models by Solainayagi *et al.* [25] and Li *et al.* [26] incorporated similar dimensions, emphasizing source *reliability* and the *credibility* of statements. These metrics represent some of the initial efforts to quantitatively assess the effectiveness of information retrieval engines in sourcing high-quality information, marking a shift from the traditional query-document relevance paradigm [27,28]. Despite their variations, these information quality metrics focus on three main common topics: i) *relevancy* (also called *usefulness* or *informativeness*) of the source to the search topic, ii) *correctness* (also called *supportiveness* or *efficacy*) of the information according to the search topic, and iii) *credibility* (also called *trustworthiness*) of the source.

Thanks to these open shared tasks, several significant methodologies have been developed to improve the search for higher-quality health information. While classical bag-of-words-based methods outperform neural network approaches in detecting health-related misinformation when training data is limited [29], more advanced approaches are needed for Web content. Specifically, research has proven the effectiveness of a hybrid approach that integrates classical handcrafted features with deep learning [18]. Further to this, multi-stage ranking systems [30,31] have been proposed, which couple the system with a label prediction model or employ T5 [33] to re-rank BM25 results. Particularly, Lima *et al.* [29] consider the stance of the search query and engage two assessors for an interactive search, integrating a continuous active learning method [58]. This approach sets a baseline of human effort in separating helpful from harmful Web content. Despite their success, these models often do not take into account the different information quality aspects in their design.

In this study, we aim to investigate the impact of multi-dimensional ranking on improving the quality of retrieved health-related information. Due to its coverage of the main information quality dimensions used in the scientific literature, we follow the empirical approach proposed in the TREC 2021 challenge, which considers *usefulness*, *supportiveness*, and *credibility* metrics, to propose a multi-dimensional ranking model. Using deep learning-based pre-trained language models [32] through transfer learning and domain adaption approaches, we categorize the retrieved Web resources according to different information quality dimensions. Specialized quality-oriented ranks obtained by re-ranking components are then fused [33] to provide the final ranked list. In contrast to prior studies, our approach relies on the automatic detection of harmful (or inaccurate) claims and uses a multi-dimensional information quality model to boost helpful resources.

The main contributions of this work are:

- We propose a multi-dimensional ranking model based on transfer learning and show that it achieves state-of-the-art in automatic, i.e., when the query stance is not provided, quality-centered ranking evaluations.
- We investigate our approach in two learning settings – transfer learning (i.e., without query relevance judgments) and domain adaptation (i.e., with query relevance judgments from a different corpus) – and demonstrate that they are capable of identifying more helpful documents than harmful ones, obtaining +5% and +7% help-harm compatibility scores, respectively.
- Lastly, we investigate how the combination of models specialized in different information dimensions impacts the quality of the results and our analysis suggests that multi-dimensional aspects are crucial for extracting high-quality information, especially for unhelpful topics.

## Methods

In this section, we introduce our search model based on multi-dimensional information quality aspects. We first describe the evaluation benchmark. Then, we detail the implementation methodology and describe our evaluation experiments using transfer learning and domain adaptation strategies.

### TREC Health Misinformation Track 2021 Benchmark

To evaluate our approach, we used the TREC Health Misinformation Track 2021 benchmark [34] organized by the National Institute of Standards and Technology (NIST) [35]. The TREC Health Misinformation Track 2021 benchmark simulates Web searches for specific health issues and interventions against a collection of English Web documents [36]. For each topic, the benchmark annotates the quality of the retrieved Web documents using a pooling approach, where the top retrieved documents by systems participating in the challenge are evaluated according to their usefulness, correctness and credibility, and subsequently labeled as helpful or harmful. In this context, helpful documents defined as those supportive of helpful treatments or that try to dissuade the reader from using unhelpful treatments while harmful documents encourage the use of unhelpful treatments or dissuade the reader from using helpful treatments [24]. See Appendix 1 Table A1 for more detail on the annotation.

### Health-related Topics

A topic in the TREC Health Misinformation Track 2021 benchmark consists of a health issue, an intervention, a query that connects the corresponding intervention to the health problem, and a description that resembles the Web search question using natural language. NIST only provided assessments for 35 of the initial 50 topics. Among the assessed topics, three were further excluded due to the absence of harmful documents. Consequently, the benchmark consisted of 32 topics: 14 labeled as helpful and 18 as unhelpful. For these queries, a total of 6,030 query-document pairs were human-annotated according to different scales of usefulness, correctness and credibility scores. A ‘helpful topic’ refers to an intervention beneficial for treating a health issue, while an ‘unhelpful topic’ indicates an ineffective intervention. The stance is supported by evidence from a credible source. Table 1 presents examples of the queries and descriptions of helpful and unhelpful topics.

**Table 1.** Examples of helpful and unhelpful topics with query and description.

| # | Query | Description | Stance |
| --- | --- | --- | --- |
| 106 | vitamin b12 sun exposure<br>vitiligo | Can vitamin b12 and sun exposure together<br>help treat vitiligo? | Helpful |
| 102 | tepid sponge bath reduce<br>fever children | Is a tepid sponge bath a good way to reduce<br>fever in children? | Unhelpful |

### Web Corpus

We used the C4 corpus, a collection of English-language Web documents sourced from the public Common Crawl web scrape [37]. The corpus comprises one billion English documents from the April 2019 snapshot. To illustrate the contradictory nature of the Web information within the corpus, in Table 2 we present two documents relevant to the topic 102 *‘tepid sponge bath reduce fever in children’*. While one article suggests against the intervention (*‘Do Not Use Sponging to Reduce a Fever’)*, another advises it could be a viable option (*‘Sponging is an option for high fevers’)*.

**Table 2.**
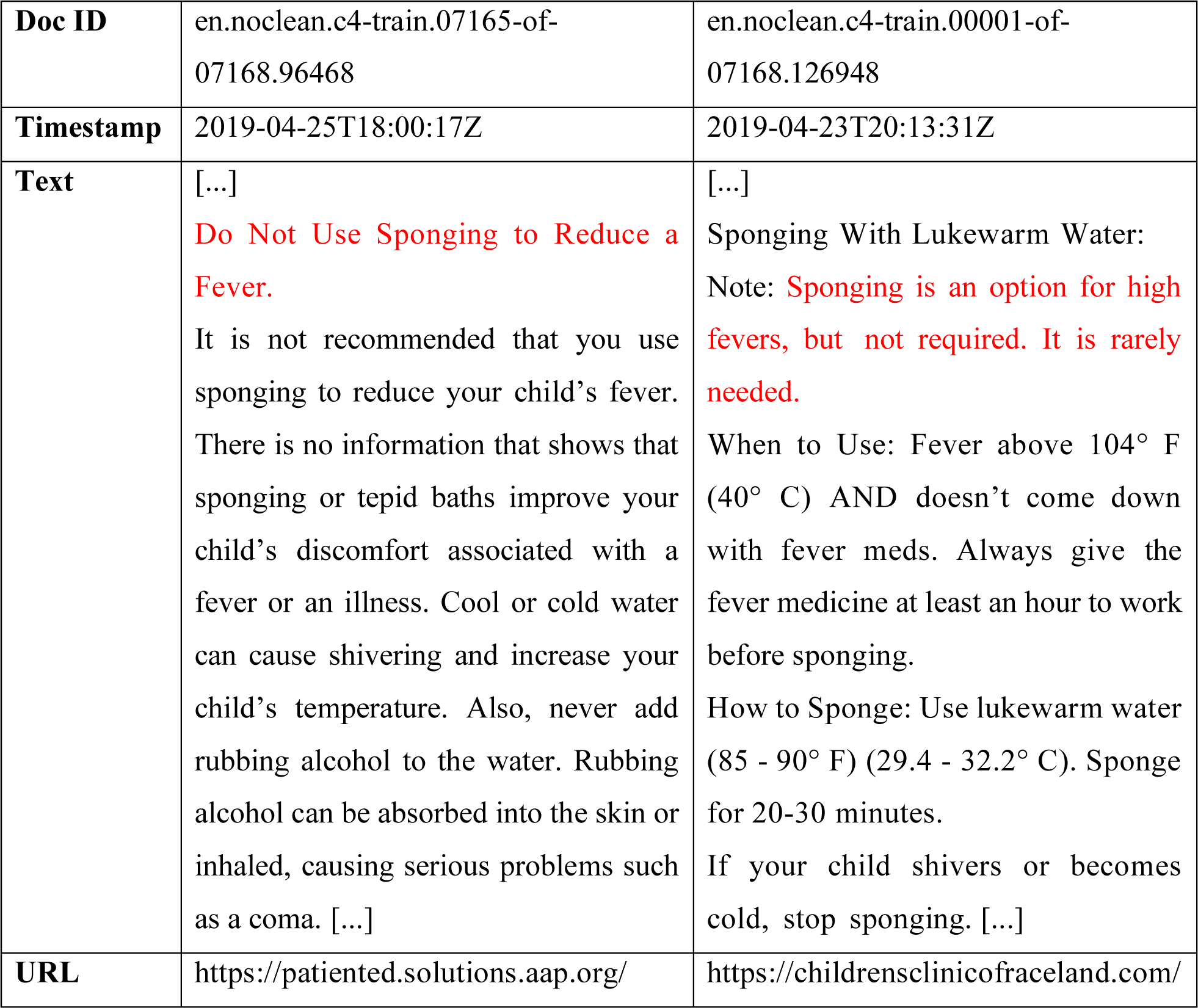
Examples of useful but contradictory documents for Topic 102 ‘Is a tepid sponge bath a good way to reduce fever in children?’.

|  |  |  |
| --- | --- | --- |
| **Doc ID** | en.noclean.c4-train.07165-of-07168.96468 | en.noclean.c4-train.00001-of-07168.126948 |
| **Timestamp** | 2019-04-25T18:00:17Z | 2019-04-23T20:13:31Z |
| **Text** | [...]<br>**Do Not Use Sponging to Reduce a Fever.**<br>It is not recommended that you use sponging to reduce your child’s fever. There is no information that shows that sponging or tepid baths improve your child’s discomfort associated with a fever or an illness. Cool or cold water can cause shivering and increase your child’s temperature. Also, never add rubbing alcohol to the water. Rubbing alcohol can be absorbed into the skin or inhaled, causing serious problems such as a coma. [...] | [...]<br>Sponging With Lukewarm Water:<br>Note: **Sponging is an option for high fevers, but not required. It is rarely needed.**<br>When to Use: Fever above 104° F (40° C) AND doesn’t come down with fever meds. Always give the fever medicine at least an hour to work before sponging.<br>How to Sponge: Use lukewarm water (85 - 90° F) (29.4 - 32.2° C). Sponge for 20-30 minutes.<br>If your child shivers or becomes cold, stop sponging. [...] |
| **URL** | https://patiented.solutions.aap.org/ | https://childrensclinicofraceland.com/ |

### Quality-based Multi-Dimensional Ranking Conceptual Model

The quality-based multi-dimensional ranking model proposed in this work is presented in Figure 1a. The information retrieval process can be divided into two phases: *preprocessing* and *multi-dimensional ranking*. In the preprocessing phase, for a given topic *j*, *N_D_* documents are retrieved based on their relevance, e.g., using a BM25 model [38]. Then, in the multi-dimensional ranking phase, we further estimate the quality of the retrieved subset of documents according to the usefulness, supportiveness, and credibility dimensions. In the following, we describe the multi-dimensional ranking approach and its implementation using transfer learning and domain adaption. Then, we describe the preprocessing step, which can be performed based on sparse or dense retrieval engines.

**Figure 1.**
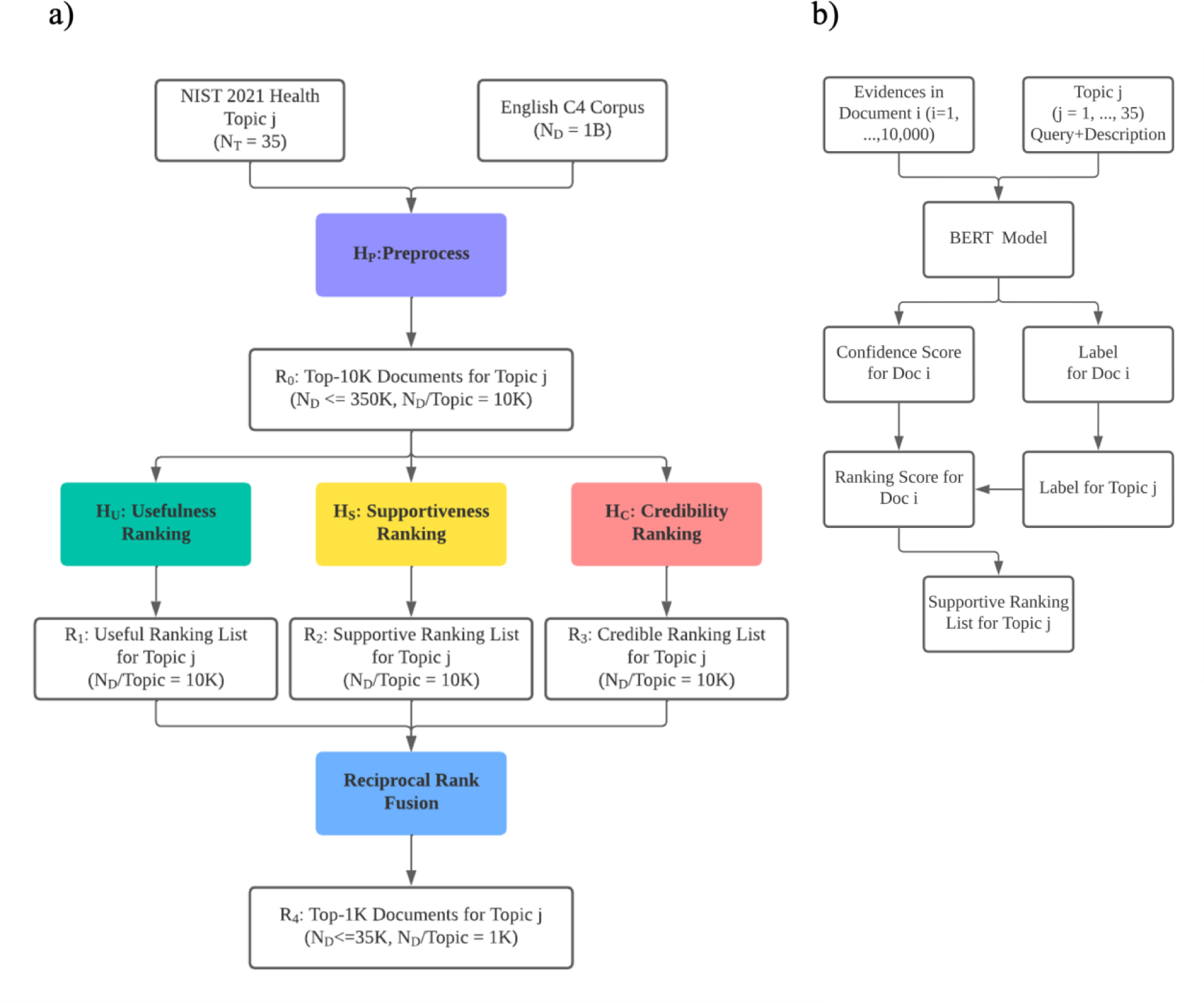
Quality-based multi-dimensional ranking models. a) General pipeline. b) Supportiveness model for the transfer learning approach.

### Multi-dimensional Ranking

To provide higher quality documents at the top ranks, we propose to use a set of machine learning models trained to classify documents according to the usefulness, supportiveness, and credibility dimensions. For the initial rank list obtained in a preprocessing phase (see details below), the documents are re-ranked in parallel according to the following strategies:

#### Usefulness

The usefulness dimension is defined as *the extent to which the document contains information that a search user would find useful in answering the topic’s question*. In this sense, it defines how pertinent a document is to a given topic. Thus, to compute the usefulness of retrieved documents, topic-document similarity models based on pre-trained language models, such as BERT-base [39], mono-BERT-large [40], and ELECTRA [41], could be used. Given a topic-document pair, the language model infers a score that gives the level of similarity between the two input text passages. While bag-of-words models, such as BM25, provide a strong baseline for usefulness, it does not consider word relations by learning context-sensitive representations as is the case with the pre-trained language models, which are used to enhance the quality of the original ranking [28].

#### Supportiveness

The supportiveness dimension defines whether *the document supports or dissuades the use of the treatment in the topic’s question*. Thus, it defines the stance of the document on the health topic. In this dimension, documents are identified under three levels: 1) *supportive*, i.e., the document supports the treatment; 2) *dissuasive*, i.e., the document refutes the treatment; and 3) *neutral*, i.e., the document does not contain enough information to make the decision [34]. To compute the supportiveness of a document to a given query, the system should be optimized so that documents that are either supportive, if the topic is helpful, or dissuasive, if the topic is unhelpful, are boosted to the top of the ranking list, which means that correct documents are boosted, and misinforming documents are downgraded.

#### Credibility

The credibility dimension defines *whether the document is considered credible by the assessor*, that is, how trustworthy is the source document. To compute this dimension, the content of the document itself could be used, e.g., leveraging language features, such as readability [42], which is assessable by the Simple Measure of Gobbledygook index [43]. Moreover, document metadata could be also employed, such as incoming and outcoming links, which can be calculated with link analysis algorithms [44], and URL addresses considering trusted sources [45], etc..

### Transfer Learning Implementation

To implement the multi-dimensional ranking model in scenarios in which relevance judgments are not available, we proposed multiple (pre-trained) models for each of the quality dimensions using transfer learning.

#### Usefulness

In this re-ranking step, we created an ensemble of pre-trained language models – BERT-base, mono-BERT-large and ELECTRA – all fine-tuned in the MS MARCO [46] dataset. Each model then predicted the similarity between the topic and the initial list of retrieved documents. Their results were finally combined using reciprocal rank fusion (RRF) [33].

#### Supportiveness

In this re-ranking step (Figure 1b), we created an ensemble of claim-checking models – RoBERTa-Large [47], BioMedRoBERTa-base [48] and SciBERT-base [49] – which were fine-tuned on the FEVER [50] and SciFact [51] datasets. Claim-checking models take a claim and a document as the information source and validate the veracity of the claim based on the document content [52]. Most claim-checking models assume that document content is ground truth. Since this is not valid in the case of Web documents, we add a further classification step, which evaluates the correctness of the retrieved documents. We use the top-*k* assignments [43] provided by the claim-checking models to define whether the topic should be supported or refuted. The underlying assumption is that a scientific fact is defined by the largest number of evidence available for a topic. Then, a higher rank is given to the correct supportive/dissuasive documents, a medium rank is given to the neutral documents and a lower rank is given to the incorrect supportive/dissuasive documents. The rank lists obtained for each model were then combined using RRF.

#### Credibility

In this step, we implemented a random forest classifier trained on the Microsoft Credibility dataset [53] with a set of credibility-related features, such as readability, openpage rank [44] and the number of cascading style sheets (CSS). The dataset manually rates 1000 Web pages with credibility scores between 1 (“very non-credible”) and 5 (“very credible”). We convert these scores for a binary classification setting – that is, the scores of 4 and 5 are considered as 1 or *credible* and scores of 1, 2, and 3 are considered as 0 or *non-credible*. For the readability score, we rely on the SMOG index [43], which estimates the years of education an average person needs to understand a piece of writing. Following Schwarz and Morris [53], we retrieve a Web page’s PageRank and use it as a feature to train the classifier. We further use the number of CSS style definitions for estimating the effort for the design of a Web page [54]. Lastly, a list of credible websites scrapped from the Health On the Net search engine [55] for the evaluated topics is combined with the baseline model to explore better performance. The result of the classifier is added to the unitary value of the Health On the Net credible sites [45].

### Domain Adaptation Implementation

To implement the multi-dimensional ranking model in scenarios in which relevance judgments are available, we compared different pre-trained language models – BERT, BioBERT [56] and BigBird [57] – for each of the quality dimensions using domain adaptation. In this case, each model is fine-tuned to predict the relevance judgment of a specific dimension, i.e., usefulness, supportiveness and credibility. While for the first two models the input size is limited to 512 tokens, BigBird allows up to 4096 tokens.

We used the TREC 2019 Decision Track [58] benchmark dataset for fine-tuning our specific quality dimension models. The TREC 2019 Decision Track benchmark dataset contains 51 topics evaluated across three dimensions: relevance, effectiveness and credibility. Adhering to the experimental design set by [59], we map the 2019 and 2021 benchmarks as follows:

- **Usefulness**: The relevance dimension (2019) was mapped to usefulness (2021), with highly relevant documents translated as very useful and relevant documents as useful.
- **Supportiveness**: The effectiveness dimension (2019) was mapped to supportiveness (2021), with effective labels reinterpreted as supportive and ineffective as dissuasive.
- **Credibility**: The credibility dimension (2019) was directly mapped to credibility (2021), using the same labels.

The 2019 track uses the ClueWeb12-B13 [60,61] corpus, which contains 50 million pages. More details on the TREC 2019 Decision Track [58] benchmark are provided in Appendix 1 Table A2. In the training phase, the language models received as input the pair topic-document and a label for each dimension according to the 2019-2021 mapping strategy. At the inference time, given a topic-document pair from TREC Health Misinformation Track 2021 benchmark, the model would infer its usefulness, supportiveness or credibility based on the dimension it was trained on.

### Preprocessing or Ranking Phase

In the preprocessing step, which is initially executed to select a short list of candidate documents for the input query, a BM25 model was used. This step was performed using a bag-of-words model due to its efficiency. Two indices were created for the C4 snapshot collection, one using standard BM25 parameters and another fine-tuned using a collection of topics automatically generated (silver standard) from a set of 4,985 indexed documents. For a given document, the silver topic was created based on the keyword2query [62] and doc2query [40] models to provide the query and description content, respectively. Then, using the silver topics and their respective documents, the *BM25* parameters of the second index were fine-tuned using grid search in a known-item search approach [63], i.e., for a given silver topic, the model should return in the top-1 the respective document used to generate it. The results of these two indices were fused using RRF.

### Evaluation Metric

We follow the official TREC evaluation strategy and employ the compatibility metric [55] to assess the performance of our models. Contrary to the classic information retrieval tasks, in which the performance metric relies on the degree of relatedness between queries and documents, in quality retrieval, harmful documents should be penalized, especially if they are relevant to the query content. In this context, the compatibility metric calculates the similarity between the actual ranking *R* provided by a model and an ideal ranking *I* as provided by the query relevance annotations. According to Equation 1, the compatibility is calculated with the Rank Biased Overlap (RBO) [64] similarity metric, which is top-weighted, with greater weight placed at higher ranks to address the indeterminate and incomplete nature of Web search results [65]:

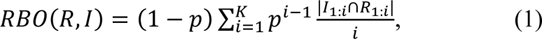

where the parameter *p* represents the searcher’s patience or persistence and is set to 0.95 in our experiments, and K is the search depth and is set to 1,000 to bring *pK*−1 as close to 0 as possible. As shown in Equation 2, an additional normalization step is added to accommodate short, truncated ideal results, so when there are fewer documents in the ideal ranking than in the actual ranking list, it does not influence the compatibility computation results:

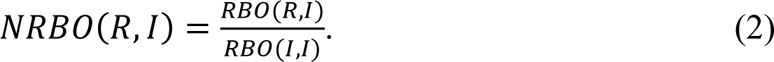

To ensure that helpful and harmful documents are treated differently, even if both might be relevant to the query content, the assessments are divided into “help compatibility” (help) and “harm compatibility” (harm) metrics. Then, to evaluate the ability of the system to separate helpful from harmful information, the “harm compatibility” results are subtracted from the “help compatibility” results, which are marked as “help-harm compatibility” (help-harm). Overall, the more a ranking is compatible with the ideal helpful ranking, the better it is. Conversely, the more a ranking is compatible with the ideal harmful ranking, the worse it is.

### Experimental Setup

The BM25 indices were created using the Elasticsearch framework (version 8.6.0). The number of documents *N_D_* retrieved per topic in the pre-processing step was set to 10,000 in our experiments. The pre-trained language models were based on open-source checkpoints from HuggingFace platform [66] and were implemented using the open-source PyTorch framework. The language models used for the usefulness dimension and their respective HuggingFace implemations are BERT base (Capreolus/bert-base-msmarco), BERT large (castorini/monobert-large-msmarco-finetune-only) and ELECTRA (Capreolus/electra-base-msmarco).. The language models used for the supportiveness dimension are RoBERTa base (allenai/biomed_roberta_base), RoBERTa large (roberta-large) and SciBERT (allenai/scibert_scivocab_uncased). For the credibility dimention, we used the random rorest algorithm of the scikit-learn library. In the domain adaptation setup, we partitioned the 2019 labeled dataset into training and validation sets, using an 80-20% split ratio, the latter is used to select the best models. Then, we fine-tuned BioBERT (dmis-lab/biobert-base-cased-v1.1) with a batch size of 16, learning rate as 1e-5, 20 epochs with early stopping set at 5 and utilizing the binary cross-entropy loss, which was optimized using the Adam optimizer. The BigBird model (google/bigbird-roberta-base) was fine-tuned with a batch size of 2, keeping all the other settings the same as the BioBERT model. All language models were fine-tuned using a single NVIDIA Tesla V100 graphics card with 32GB of memory. Results are reported using the compatibility and normalized discounted cumulative gain (nDCG) metrics. For reference, they are compared to the results of other participants of the official TREC Health Misinformation 2021 track, which have submitted runs for the automatic evaluation, i.e., without using information about the topic stance. The code repository is available at [67].

## Results

In Table 3, we present the performance results of our quality-based retrieval models using the TREC Health Misinformation 2021 benchmark. Helpful compatibility (help) considers only helpful documents of the relevant judgment, while harmful compatibility (harm) considers only harmful documents, and help-harm their compatibility difference (see Appendix 1-Table A1 for further detail). Additionally, we show the nDCG scores calculated using helpful (help) documents or harmful (harm) documents of the relevant judgment. The ‘helpful*T*’, ‘unhelpful*T*’ and the ‘all*T*’ denote helpful topics, unhelpful topics and all topics, respectively. *HU*, *HS* and *HC* rankings represent the combination of the preprocessing (*HP*) results with and the re-rankings results for usefulness (*HU’*), supportiveness (*HS’*) and credibility (*HC’*), respectively. For reference, we show our results compared to the models participating in the TREC Health Misinformation Track 2021: **Pradeep *et al*.** [30] used the default BM25 ranker from Pyserini. Their re-ranking process incorporated a mix of mono and duo T5 models as well as Vera [68] on different topic fields.

**Table 3.** Performance results for the quality-based retrieval models. *H_U_*: Usefulness model; *H_S_*: Supportiveness model; *H_C_*: Credibility model. Help: results considering only helpful documents in the relevance judgment. Harm: results considering only harmful documents in the relevance judgment. all*T*: all topics; helpful*T*: helpful topics; unhelpful*T*: unhelpful topics.

| Model | nDCG |  | Compatibility |  |  |  |  |
| --- | --- | --- | --- | --- | --- | --- | --- |
| | Help ↑ | Harm ↓ | Help ↑ | Harm ↓ | Help-Harm ↑ | | |
| | $all_T$ | $all_T$ | $all_T$ | $all_T$ | $helpful_T$ | $unhelpful_T$ | $all_T$ |
| BM25 [38] | 0.516 | 0.360 | 0.122 | 0.144 | 0.158 | -0.162 | -0.022 |
| Pradeep *et al.* [31] | 0.602 | 0.378 | **0.195** | 0.153 | **0.234** | -0.106 | 0.043 |
| Abualsaud *et al.* [69] | 0.302 | **0.185** | 0.164 | 0.123 | 0.179 | -0.067 | 0.040 |
| Schlicht *et al.* [70] | 0.438 | 0.309 | 0.121 | 0.103 | 0.157 | -0.089 | 0.018 |
| Pichel *et al.* [71] | **0.603** | 0.363 | 0.163 | 0.155 | 0.163 | -0.113 | 0.008 |
| Bondarenko *et al.* [73] | 0.266 | 0.226 | 0.129 | 0.144 | 0.150 | -0.144 | -0.015 |
| Transfer Learning |  |  |  |  |  |  |  |
| $H_U$ | <u>0.538</u> | 0.324 | <u>0.142</u> | **0.087** | 0.156 | **-0.022** | **0.056** |
| $H_U + H_S$ | 0.477 | <u>0.315</u> | 0.130 | 0.092 | 0.151 | -0.049 | 0.038 |
| $H_U + H_S + H_C$ | 0.484 | 0.320 | 0.137 | 0.095 | <u>0.169</u> | -0.057 | 0.042 |
| Domain Adaptation |  |  |  |  |  |  |  |
| $H_U$ | 0.510 | 0.327 | 0.128 | 0.100 | 0.146 | -0.063 | 0.029 |
| $H_U + H_S$ | 0.482 | 0.319 | 0.108 | 0.089 | 0.108 | -0.050 | 0.019 |
| $H_U + H_S + H_C$ | 0.502 | 0.325 | 0.131 | 0.094 | 0.147 | -0.048 | 0.037 |

**Abualsaud *et al***. [69] created filtered collections that focus on filtering out non-medical and unreliable documents, which were then used for retrieval with Anserini’s BM25.

**Schlicht *et al.*** [70] also used Pyserini’s BM25 ranker and Bio Sentence BERT to estimate usefulness and RoBERTa for credibility. The final score was a fusion of these individual rankings. **Pichel *et al***. [71] employed BM25 and RoBERTa for re-ranking and similarity assessment of top 100 documents, trained an additional reliability classifier, and merged scores using CombSUM [72] or Borda Count.

**Bondarenko *et al*.** [73] used Anserini’s BM25 and PyGaggle’s MonoT5 for two baseline rankings, then re-ranked the top 20 from each using three argumentative axioms on seemingly argumentative queries.

Our approach provides state-of-the-art results for automatic ranking systems in the transfer learning setting, with help-harm compatibility of +5.6%. This result is obtained by the usefulness model (*HU*), which is the combination of preprocessing and usefulness re-ranking. It outperforms the default BM25 model [38] by 7% (*p*-value = .04) and the best automatic model from the TREC 2021 benchmark (Pradeep *et al*. [31]) by 1%. In this case, while the help and harm compatibility metrics individually exhibit statistical significance (*p*-value = .02 and *p*-value = .01 respectively), the improvement in help-harm compatibility compared to the best automatic model is not statistically significant (*p*-value = .7). The usefulness model also stands out by achieving the best help and harm compatibility among our models (14.2% and 8.7%, respectively; *p*-value = .5). Notice that for the latter metric, the closest to 0, the best is the performance. Interestingly, the usefulness model attains the highest nDCG score on help for all topics as well (*p*-value = .03). The combination of usefulness, supportiveness, and credibility models (**H_U_*+ *H_S_* + *H_C_**) provides the best help-harm (+16.9%) for helpful topics among our models (**H_U_**: *p*-value = .4; **H_U_* + *H_S_**: *p*-value = .04).

Meanwhile, when calculating nDCG scores on harm, the combination of usefulness and supportiveness model (*H_U_* + *H_S_*) in the transfer learning and domain adaption settings outperforms the other model combinations (*p*-value = .5), indicating a different perspective of the best-performing model. Lastly, differently from what would be expected, in the domain adaption setting, the performance was poorer as compared to the simpler transfer learning approach (2% decrease on average for the compatibility metric; *p*-value = .02).

### Performance Stratification by Quality Dimension

In Table 4, we show the help, harm and help-harm compatibility scores on the individual quality-based re-ranking models, which disregarding the preprocessing step (prime index). Additionally, we provide the nDCG scores for a more comprehensive view of the models’ performance. The *HP* represents the preprocessing, and *HU’, HS’,* and *HC’* stand for re-rankings for usefulness, supportiveness and credibility, respectively.

**Table 4.** Performance results for the individual ranking models. *Hp*: Preprocess; *HU’*: Usefulness model; *HS’*: Supportiveness model; *HC’*: Credibility model. *HU’*, *HS’* and *HC’* rankings are not combined with *Hp* as opposed to *HU*, *HS* and *HC.* Help: results considering only helpful documents in the relevance judgment. Harm: results considering only harmful documents in the relevance judgment. all*T*: all topics; helpful*T*: helpful topics; unhelpful*T*: unhelpful topics.

| Setting | Model | nDCG |  | Compatibility |  |  |  |  |
| --- | --- | --- | --- | --- | --- | --- | --- | --- |
|  |  | Help ↑ | Harm ↓ | Help ↑ | Harm ↓ | Help-Harm ↑ |  |  |
|  |  | all<sub>T</sub> | all<sub>T</sub> | all<sub>T</sub> | all<sub>T</sub> | helpful<sub>T</sub> | unhelpful<sub>T</sub> | all<sub>T</sub> |
| | $H_p$ | **0.538** | 0.341 | **0.126** | 0.111 | **0.127** | -0.072 | 0.015 |
| Transfer Learning | $H_U'$ | 0.438 | 0.264 | 0.115 | 0.080 | 0.106 | -0.020 | 0.036 |
| | $H_S'$ | 0.140 | **0.102** | 0.026 | 0.024 | 0.021 | -0.013 | 0.002 |
| | $H_C'$ | 0.131 | 0.113 | 0.031 | 0.035 | 0.033 | -0.032 | -0.003 |
| Domain Adaptation | $H_U'$ | 0.436 | 0.277 | 0.077 | 0.038 | 0.099 | -0.008 | **0.039** |
| | $H_S'$ | 0.368 | 0.251 | 0.030 | **0.015** | 0.030 | **0.003** | 0.014 |
| | $H_C'$ | 0.443 | 0.296 | 0.079 | 0.064 | 0.104 | -0.055 | 0.014 |

In the transfer learning setting, the usefulness model (*HU’*) achieves the highest help-harm compatibility (+3.6%; *p*-value = .2). The preprocessing model gives the best help compatibility (+12.7%; *HU’*: *p*-value = .7; *HS’* and *HC’*: *p*-value < .001). Additionally, the preprocessing model yields the highest nDCG score for help (*HU’*: *p*-value = .1; *HS’* and *HC’*: *p*-value < .001). On the other hand, the preprocessing model shows the highest harm compatibility (+11.1%; *HU’*: *p*-value = .33; *HS’* and *HC’*: *p*-value < .01). The combination of the preprocessing and usefulness models (i.e., *HU* = +5.6%) improves the preprocessing model by 4.1% (from +1.5% to +5.6% on the help-harm compatibility; *p*-value = .06). For harm compatibility, the supportiveness model (*HS’*) achieves the best performance among the individual models (+2.4%; *Hp*: *p*-value = < .001; *Hu’*: *p*-value = .03; *HC’*: *p*-value = .34; ).

In the domain adaptation setting, the usefulness model (*HU’*) reaches help-harm compatibility of +3.9%, similarly outperforming the other models (*p*-value = .32). The supportiveness model (*HS’*) achieves the best performance on harm compatibility (+1.5%; *p*-value = .07) and on help-harm compatibility for unhelpful topics (+0.3%; *p*-value = .5). Notice that +0.3% is the only positive help-harm compatibility for harmful topics throughout all the individual and combined models on both settings including the preprocessing step. Lastly, in the domain adaption setting, the performance of individual models was better as compared to the simpler transfer learning approach (1% increase on average for the compatibility metric; *p*-value = .19).

### Re-ranking for the Top-N Documents

To further illustrate the effectiveness of supportiveness and credibility dimensions, in Figure 2 we re-rank only the top-n documents using the results of the usefulness model (*HU*) as the basis. As we can notice from **Table 4**, the overall effectiveness of the supportiveness (*HS’*) and credibility (*HC’*) models are considerably lower compared to the usefulness (*HU’*) model. The reason is that the relevance judgments are created using a hierarchical approach: only useful documents are further considered for supportiveness and credibility evaluations. As we re-rank the documents in supportiveness and credibility dimensions without taking this hierarchy into account, their results might not be optimal. For example, low-ranking documents (i.e., not useful) could have high credibility and during the re-ranking process be boosted to the top ranks. Thus, to the usefulness model (*HU*) results, we apply the supportiveness (*HS’*) and credibility (*HC’*) models for re-ranking the top 10, 20, 50, 100 and 1000 documents, obtaining two new rankings, which are combined using RRF.

**Figure 2.**
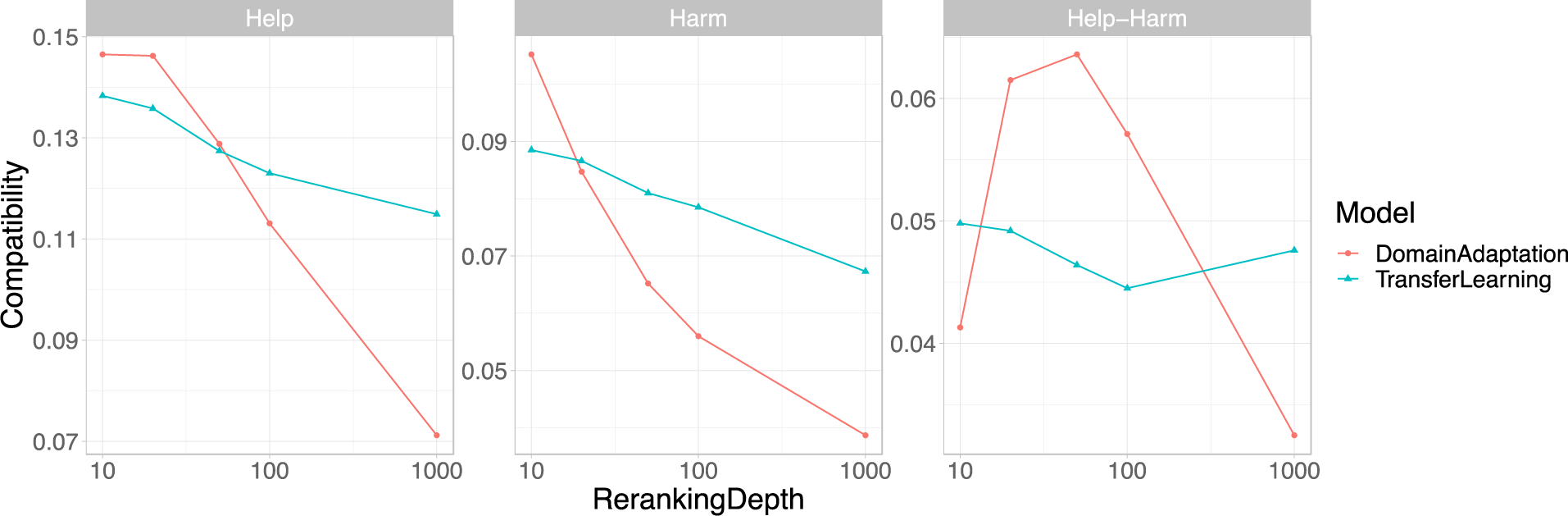
Help, harm, and help-harm compatibility performance for the top 10, 20, 50, 100 and 1000 re-ranking depths taking the results of the usefulness (HU) as the basis.

As the reranking depth increases from 10 to 1000, we observe a decrease in both help and harm compatibility. This suggests that both helpful and harmful documents are downgraded due to the inclusion of less useful but potentially supportive or credible documents. In the transfer learning setting, as the reranking depth increases, the help-harm compatibility decreases until the depth reaches 100. Beyond this point, we observe a slight increase at the depth of 1000. In the domain adaptation setting, the help-harm compatibility increases above +6% when the reranking depth is between 20 and 50. This implies that, following the procedure of human annotation, by considering only the more useful documents, the supportiveness and credibility dimensions can help to retrieve more helpful than harmful documents.

### Quality Control

One of the advantages of the proposed multi-dimensional model is that we can optimize the results according to different quality metrics. In Figure 3, we show how the compatibility performance varies by changing the weight of specific models: *H_P_*, *H_U_′*, *H_S_′*, and *H_C_′*. We normalize the score of the individual models to the unit and combine them linearly using a weight for one model between 0 and 2, while fixing the weight for the other three models at 0.33. For example, to see the influence of *H_P_*in the final performance, we fix the weights of *H_U_′*, *H_S_′*, and *H_C_′* at 0.33 and vary the weight of *H_P_* between 0 and 2. With weight 0, the reference model does not account for the final rank, while with weight 2, its impact is twice the sum of the other three models.

**Figure 3.**
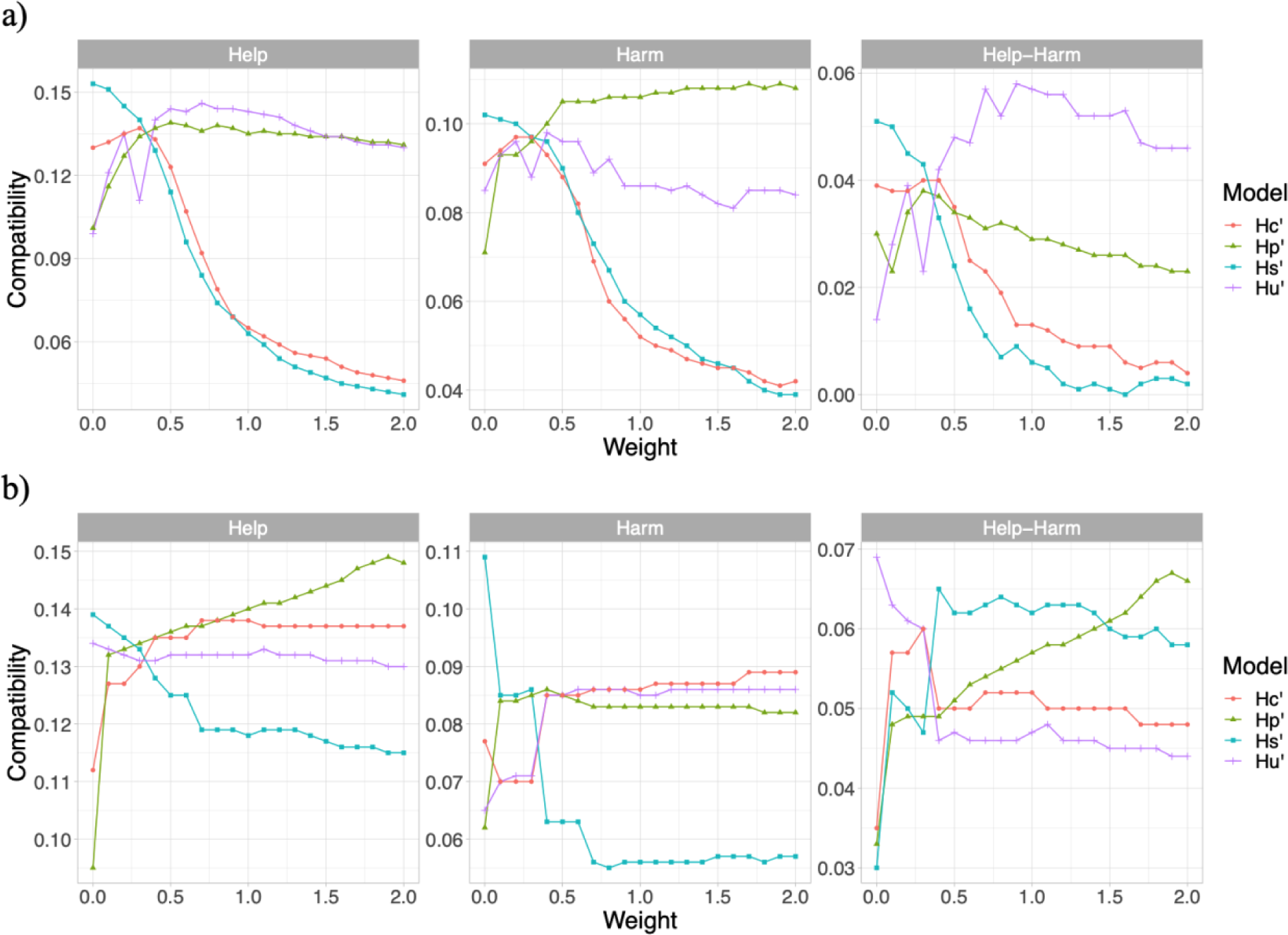
Help, harm, and help-harm compatibility with weights added to specific models *H_P_*, **H_U_*^′^*, **H_S_* ^′^* and **H_C_*^′^*. a) Transfer learning approach. b) Domain adaptation approach.

In the transfer learning setting, when we increase the weight of preprocessing and usefulness models, the help-harm compatibility increases to the best performance (+4.1% and +5.6%) and then decreases slightly. For the supportiveness and credibility dimensions, the help-harm compatibility begins to decrease once the weight is added. These results imply that the compatibility decreases with the weight addition regardless of whether it is helpful compatibility, harmful compatibility, or the difference between the two.

In the domain adaptation setting, when we increase the weight of preprocessing, supportiveness, and credibility models individually, the help-harm compatibility increase and then converge to +6.6%, +5.9%, and +4.8%, respectively. For the usefulness model, the help-harm compatibility decreases once the weight is added until it converges to +4.4%. It is worth noticing that, by combining the rankings linearly, the help-harm compatibility obtained from the domain adaptation setting may exceed the results we obtained when performing ranking combination with RRF (+3.7%), as well as the state-of-the-art result (+5.6%) in the transfer learning setting. The highest help-harm compatibility scores of each weighting combination are +6.6%, +6.8%, +6.5%, and +5.9%, when varying the weights of *H_P_*, *H_U_′*, *H_S_′*, and *H_C_′*, respectively.

### Model Interpretation

To semantically explain the variation of help-harm compatibility, we set the search depth *K* to 10. The help, harm, and help-harm compatibility of the three models are shown in *Table 5*. The help-harm compatibility is 1 when only helpful documents are retrieved in the top 10. Conversely, the help-harm compatibility is -1 when only harmful documents are retrieved in the top 10. A variation of 10% in the help or harm compatibility corresponds roughly to one helpful document exceeding the number of harmful documents retrieved in the top 10. Overall, the results show that retrieving relevant documents for health-related queries is hard, as on average only 1.5 out of 10 documents are relevant (helpful or harmful) to the topic. In addition, we interpret that the three models retrieve on average twice the number of helpful documents as compared to harmful documents. Particularly, *H_U_* has on average around one more helpful than harmful document in the top 10, out of the 1.5 relevant retrieved. We also present the same analysis result for the domain adaptation setting, which also implies that, when the rankings are combined with RRF, the transfer learning approach outperforms the domain adaptation approach.

**Table 5.** Help, harm, and help-harm compatibility with search depth set to 10 for the transfer learning setting and the domain adaptation setting.

| Setting | Model | Help ↑ | Harm ↓ | Help-Harm ↑ |
| --- | --- | --- | --- | --- |
| Transfer Learning | $H_U$ | **0.112** | **0.047** | **0.065** |
| | $H_U + H_s$ | 0.088 | 0.050 | 0.038 |
| | $H_U + H_s + H_c$ | 0.099 | 0.056 | 0.044 |
| Domain Adaptation | $H_U$ | 0.094 | 0.060 | 0.034 |
| | $H_U + H_s$ | 0.074 | 0.070 | 0.003 |
| | $H_U + H_s + H_c$ | 0.087 | 0.076 | 0.011 |

## Discussion

We propose a quality-based multi-dimensional ranking model to enhance the usefulness, supportiveness, and credibility of retrieved Web resources for health-related queries. By adapting our approach in a transfer learning setting, we show state-of-the-art results in the automatic quality ranking evaluation benchmark. We further explore the pipeline in a domain adaptation setting and show that in both settings, the proposed method can identify more helpful than harmful documents measured by +5% and +7% help-harm compatibility scores, respectively. By combining different re-ranking strategies, we show that multi-dimensional aspects have a significant impact on retrieving high-quality information, particularly for unhelpful topics.

The quality of Web documents is biased in terms of topic stance. For all models, helpful topics achieve higher help compatibility, while unhelpful topics achieve higher harm compatibility. The implication is that Web documents centered around helpful topics are more likely to support the intervention and are helpful. On the other hand, Web documents focusing on unhelpful topics present an equal chance of being supportive or dissuasive on the intervention and are helpful or harmful. Among other consequences, if Web data is used to train large language models without meticulously crafted training examples using effective dataset search methods [74], as the one proposed here, they are likely to further propagate health misinformation.

Automatic retrieval systems tend to find more helpful information on helpful topics with the information biased towards helpfulness and find more harmful information on unhelpful topics with the information slightly biased towards harmfulness. The help-harm compatibility ranges from +2.3% to +15.3% for helpful topics and -5.7% to +0.2% for unhelpful topics. The difference shows that for the improvement of quality-centered retrieval models, it is especially important to focus on unhelpful topics. Moreover, while specialized models might provide enhanced effectiveness, their combination is not straightforward. In our experiments, we show that supportiveness and credibility models should be applied only in the top 20 to 50 retrieved documents to achieve optimal performance.

Finding the correct stance automatically is another key component of the automatic model. Automatic models show the ability to prioritize helpful documents, resulting in positive help-harm compatibility. However, they are still far from state-of-the-art manual models, with help-harm compatibility scores ranging from +20.8% [69] to +25.9% [31]. We acknowledge that the help-harm compatibility can improve significantly with the correct stance given. This information is nevertheless unavailable in standard search environments and thus the scenario analyzed in this work is more adapted to real-world applications.

This work has certain limitations. In the domain adaptation setting, we simplified the task to consider two classes within each dimension for the classification due to the limited variety available in the labeled dataset. Alternatively, we could add other classes from documents that have been retrieved. Moreover, the number of topics used to evaluate our models is limited (n=32), despite including 6’030 human-annotated query-document pairs, and thus reflects only a small portion of misinformation use cases.

To conclude, the proliferation of health misinformation in Web resources has led to mistrust and confusion among online health advice seekers. Automatic maintenance of factual discretion in Web search results is the need of the hour. We propose a multi-dimensional information quality ranking model, which utilizes usefulness, supportiveness, and credibility to strengthen the factual reliability of the health advice search results. Experiments conducted on publicly available datasets show that the proposed model is promising, achieving state-of-the-art performance for automatic ranking in comparison to various baselines implemented on the TREC Health Misinformation 2021 benchmark. Thus, the proposed approach could be used to improve online health searches and provide quality enhanced information for health information seekers. Future research could explore more fine-grained classification models for each dimension and a model simplification could provide an advantage for real-world implementations.

## Acknowledgments

Innosuisse project funding numbers 55441.1 IP-ICT and 101.466 IP-ICT.

## Data Availability

The data sets generated during and/or analyzed during this study are available in the TREC Health Misinformation Track repository [75] and GitLab repository [67].

## Author contributions statement

BZ, NN, and DT prepared the data, conceived, and conducted the experiments, as well as analyzed the results. BZ, NN, and DT drafted the manuscript. All authors reviewed the manuscript.

## Abbreviations

BERT: Bidirectional Encoder Representations from Transformers
BM25: Okapi Best Match 25
CSS: cascading style sheets
CLEF: Conference and Labs of the Evaluation Forum
C4: Colossal, Cleaned Crawl Corpus
NIST: National Institute of Standards and Technology
NLP: natural language processing
RBO: Rank Biased Overlap
SMOG: Simple Measure of Gobbledygook
TREC: Text REtrieval Conference

## Appendices

### 1 Additional Information on Benchmark Datasets

**Table A1.**
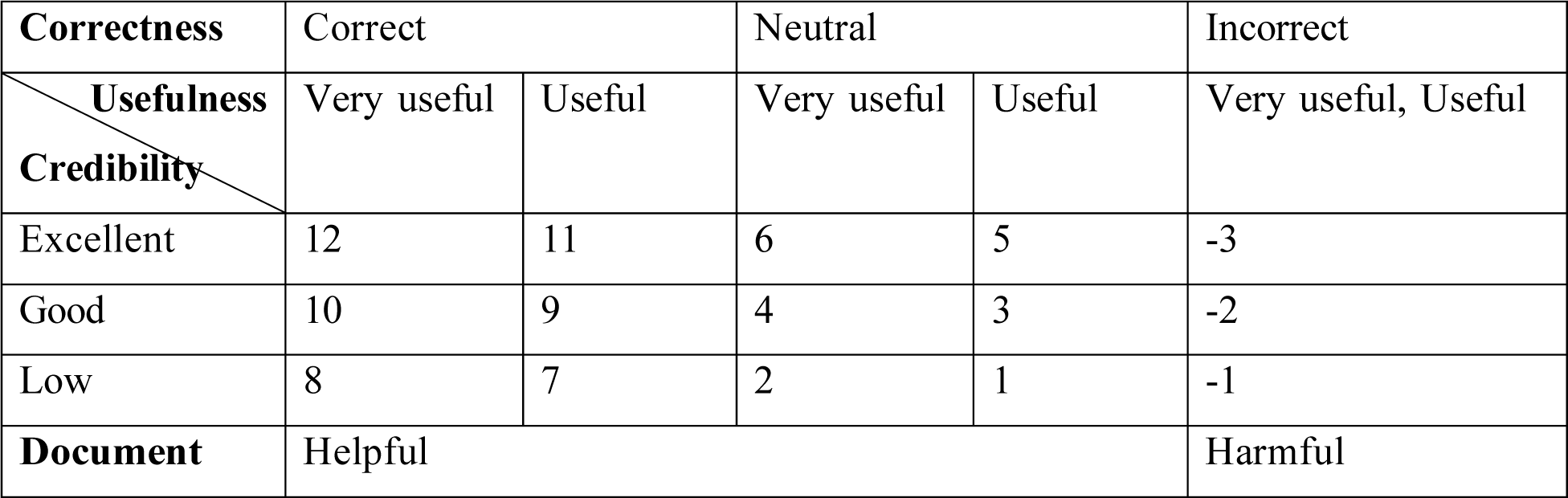
Dimensions for assessing topic-Web document pairs and their respective assessment score.

| **Correctness** | Correct |  | Neutral |  | Incorrect |
| --- | --- | --- | --- | --- | --- |
| **Usefulness**<br>**Credibility** | Very useful | Useful | Very useful | Useful | Very useful, Useful |
| Excellent | 12 | 11 | 6 | 5 | -3 |
| Good | 10 | 9 | 4 | 3 | -2 |
| Low | 8 | 7 | 2 | 1 | -1 |
| **Document** | Helpful |  |  |  | Harmful |

**Table A2.** Mappings of Labeled Data for Training/Validation (2019) and Test (2021).

| **Dataset** | **Usefulness** |  | **Supportiveness** |  | **Credibility** |  |
| --- | --- | --- | --- | --- | --- | --- |
|  | Very Useful | Useful | Support | Refute | Credible | Not Credible |
| Train (2019) | 826 | 1832 | 2572 | 86 | 1417 | 1238 |
| Dev (2019) | 113 | 414 | 452 | 75 | 315 | 212 |
| Test (2021) | 2089 | 2467 | 3667 | 889 | 2831 | 1706 |

### 2 Fine-Tuning in the Domain Adaptation Setting

#### Statement Generation

We convert the query from a question to a statement format. We tokenize the query using the SpaCy scispacy library (en_core_sci_sm) and extract name entities. Each entity is treated as a single token and the first two tokens are swapped if the initial token is a question word. For instance, the query ‘Is a tepid sponge bath a good way to reduce fever in children?’ is translated automatically into ‘a tepid sponge bath a good way to reduce fever in children.’.

#### Sentence Selection

Given the maximum token limit at 512 of a BERT model, with the same model as the encoder, we calculate the cosine similarity between the statement and sentence representations. We rank sentences by their relevance to the statement and extract the most significant text compiled until reaching the token limit of 512. For BioBERT, we implement two methods: truncating retrieved documents to 512 tokens (TD) and using sentence selection (SS).

#### Ranking Strategies

**Table B1:**
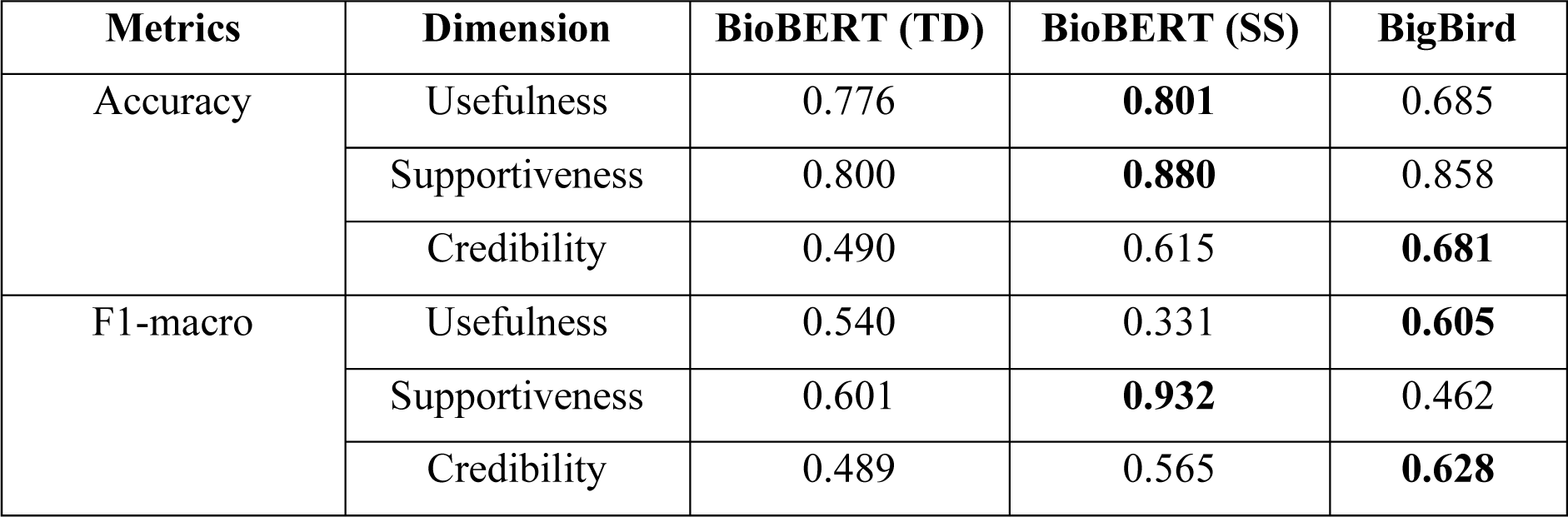
Classification accuracy and F1 score on the validation set. (TD: truncating retrieved documents; SS: sentence selection; Note: Data in this table is compared row-wise.)

Based on the F1 Marco scores in Table B1, we select the best-performing model for each dimension to re-rank the preprocessing ranking list. For usefulness and credibility, the positive confidence score is used when the predicted label is very useful or credible, while the negative score is used when the predicted label is useful or not credible. For supportiveness, we utilize the top-10 most credible documents to determine the stance of the topic. If the stance of the topic matches the stance of the document, we use the positive confidence score, otherwise we use the negative confidence score. We employ these updated confidence scores to re-rank the documents for each dimension.

### 3 Supporting Experiment Results

**Table C1:** nDCG usefulness, correctness, and credibility across quality-based retrieval models.

| Setting | Model | nDCG |  |  |
| --- | --- | --- | --- | --- |
|  |  | Useful | Correct | Credible |
| Transfer Learning | $H_U$ | 0.595 | **0.596** | 0.514 |
| | $H_U + H_S$ | 0.559 | 0.546 | 0.468 |
| | $H_U + H_S + H_C$ | 0.564 | 0.557 | 0.486 |
| Domain Adaptation | $H_U$ | **0.607** | 0.566 | 0.547 |
| | $H_U + H_S$ | 0.578 | 0.540 | 0.529 |
| | $H_U + H_S + H_C$ | 0.586 | 0.563 | **0.563** |

We use nDCG as a metric to provide another perspective on evaluating usefulness, correctness, and credibility. The usefulness model (*HU*) in the transfer learning setting shows the highest nDCG for correctness (0.5*96*), which implies that correctness can already be leveraged by improving only the usefulness dimension of the model. In the domain adaptation setting, the usefulness model (*HU*) demonstrates the highest nDCG for usefulness (0.607). The combination of usefulness, supportiveness, and credibility models (*H_U_* + *H_S_* + *H_C_*) presents the highest nDCG for credibility (0.563). The results imply that the usefulness and credibility dimensions each effectively contribute to their respective attributes in the retrieved documents.

In Figure C1, we show the average compatibility for all the topics as the search depth K varies. In the transfer learning setting, when the search depth is 1, we compare the top-1 document in the ranking list and the query relevance set. In this case, the compatibility is either 0 or 1 for each topic. When the search depth is above 1, we compare the top-K documents and the compatibility for each topic varies between 0 and 1. The compatibilities increase almost monotonically until the search depth K becomes 100 as the query reaches the average number of documents in the relevance set (i.e., 92 documents) and subsequently becomes saturated. Interestingly, we observe that for help-harm compatibility, the model *H_U_* spikes at the depth of 3 due to the higher proportion of helpful documents retrieved as compared to harmful documents. Further, with the depth increase, we notice that the difference in proportions of helpful and harmful documents diminishes steeply between depths 3 to 10. The results in the domain adaptation setting indicate the same trend.

**Figure C1.**
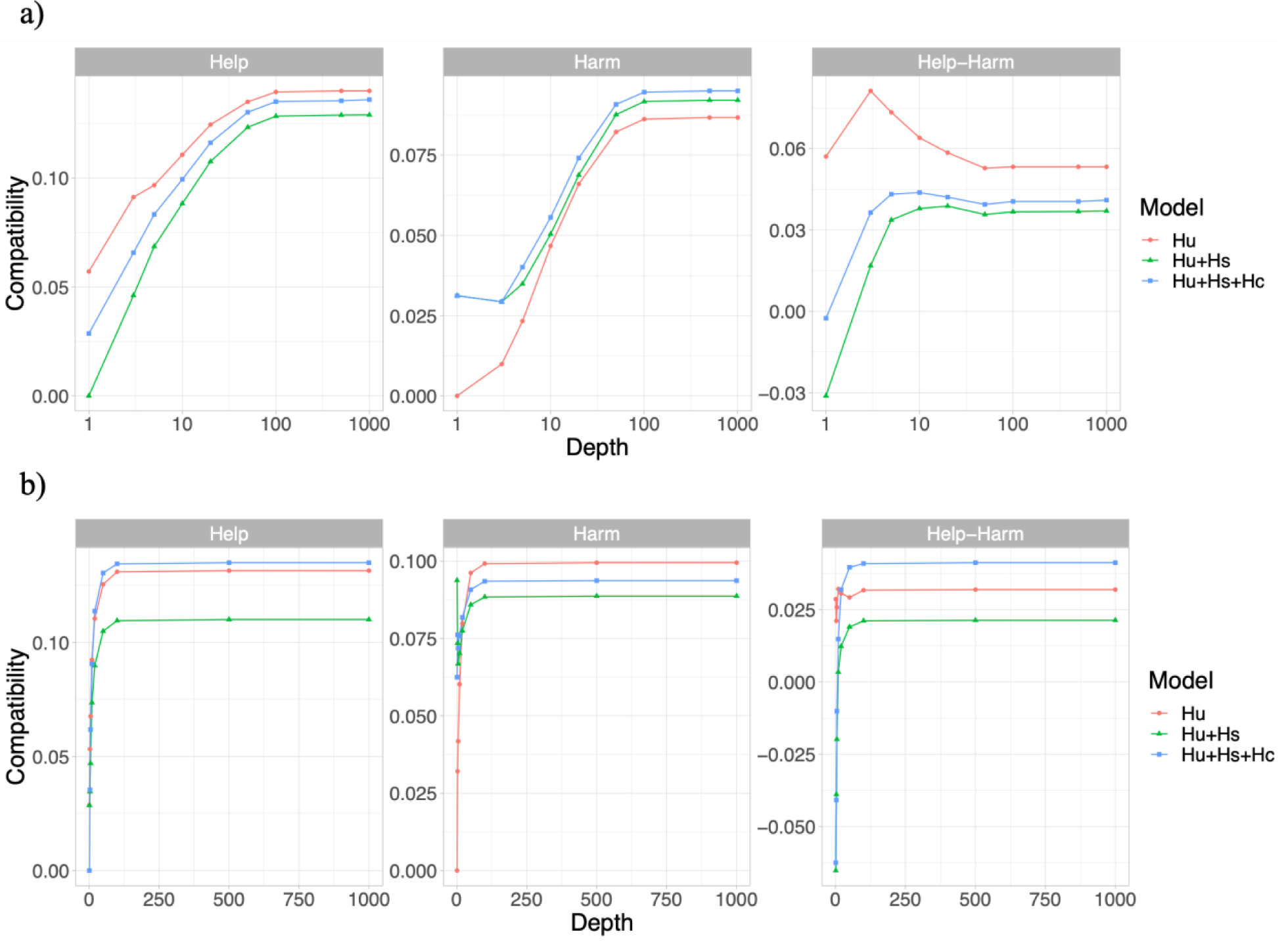
Help, harm, and help-harm compatibility with different search depths. a) Transfer learning approach. b) Domain adaptation approach.

